# Effect of an Electronic Health Record–Integrated Clinical Dashboard for Radiologists on STAT Priority Chest Radiograph Reporting and Downstream Care: A Stepped-Wedge Cluster Randomized Trial

**DOI:** 10.64898/2026.09.23.26363355

**Authors:** Jason D. Balkman, Sahithi R. Chimmula, Allison M. Lam, Joshua R. Nugent, Tejomay Gadgil, Andrew L. Avins, Dana R. Sax, Vignesh A. Arasu

**Author notes:** **Corresponding author:** Jason D. Balkman, MD, Department of Radiology, Kaiser Permanente, Pleasanton, CA. **Trial registration:** ClinicalTrials.gov NCT06785246. Institutional study ID 2142507.

## Abstract

**Background:** Chest radiographs (CXRs) are the most common imaging exam performed, but CXR reports can be nonspecific due to incomplete history, relying on vague terms like “opacities” to convey diagnostic uncertainty. We prospectively evaluated whether introducing a clinical dashboard with relevant patient information affects CXR report specificity and efficiency, report concordance with discharge diagnoses, and follow-up chest CT utilization.

**Methods:** In this stepped-wedge cluster randomized trial (NCT06785246), a clinical dashboard for radiologists was rolled out with user education sessions across 15 service areas of an integrated health system from June 2024 to June 2025. Emergency department (ED) and inpatient CXRs were included. A large language model (GPT-4o) was validated using a Delphi panel of three experts to classify report specificity and CXR report diagnosis. A subsequent model release (GPT-5.4) graded each CXR report impression as negative, specific abnormal, or nonspecific abnormal. It also judged whether an abnormal impression matched an ED discharge diagnosis. The primary outcome was the proportion of specific reports (negative or specific abnormal). Secondary outcomes were report–discharge diagnosis concordance, follow-up chest CT, antibiotic and diuretic orders, and CXR reporting time. Mixed-effects models adjusted for time, patient age, and radiograph view, with random effects for radiologists, facilities, and service area.

**Results:** The trial included 404,860 ED and inpatient encounter STAT priority CXRs read by 387 radiologists (256,303 control; 148,557 intervention). The adjusted proportion of specific reports was 82.2% in control periods and 82.1% in intervention periods (difference, −0.10 percentage points; 95% CI, −0.53 to 0.34; P=.66). Among abnormal reports, concordance with a discharge diagnosis rose from 40.9% to 42.2% (difference, 1.24 points; 95% CI, 0.20 to 2.28; P=.02). Follow-up chest CT fell from 11.4% to 10.9% (difference, −0.55 points; 95% CI, −0.92 to −0.17; P=.004). Antibiotic orders (24.0% vs 23.9%) and diuretic orders (14.7% vs 14.8%) within 7 days did not change, nor did reporting time (difference, −0.8 seconds; 95% CI, −2.7 to 1.0).

**Conclusions:** Giving radiologists EHR context through a dashboard did not change the specificity of STAT priority CXR reports or prescribing patterns. It was associated with small improvements in diagnostic concordance and follow-up CT use, without slowing reporting. Validated large language models make it feasible to measure report quality across a health system.

**Trial registration:** ClinicalTrials.gov NCT06785246.

## Introduction

The chest radiograph (CXR) is the most frequently performed imaging examination. In Kaiser Permanente Northern California (KPNC), CXRs account for nearly one third of imaging volume, including about 500,000 emergency department (ED) and inpatient studies each year. Despite this volume, the diagnostic yield of a CXR is often limited. Part of the limitation is technical.

Different diseases produce similar findings, and CXR agrees poorly with CT for detecting pulmonary opacities.^1^ Another limitation is that radiologists usually see only a brief indication on the order, and that indication may be incomplete or discordant with the chart.^2^ Clinical information improves the accuracy, confidence, and clinical relevance of image interpretation^3,4^, but high volumes and turnaround-time pressure leave little time to search the chart.

Limited context encourages nonspecific reporting. A radiologist who cannot distinguish pneumonia from pulmonary edema on the image alone may report “bilateral opacities” or offer a differential (“edema versus infection”). Expressions of uncertainty in radiology reports are interpreted inconsistently by referring clinicians.^5,6^ A nonspecific report may prompt additional testing, such as chest CT, or parallel treatment with both antibiotics and diuretics. Simple clinical data can resolve some of this uncertainty: a normal B-type natriuretic peptide (BNP) argues strongly against heart failure^7,8^, and fever or leukocytosis supports infection.

EHR-driven radiology workflows have been proposed as a way to bring such data to the reading room^9^, but effects of radiology clinical dashboards on reporting and patient care have not been tested in a randomized design. We developed Grasshopper, a web dashboard that opens alongside the radiology report and displays laboratory values, vital signs, and clinical notes relevant to the examination being read. We conducted a stepped-wedge cluster randomized trial to test whether rollout of the dashboard with radiologist education (1) increased the specificity of STAT priority (ED and inpatient) CXR reports and (2) changed report concordance with the clinical diagnosis, follow-up chest CT use, and antibiotic and diuretic prescribing. Because the trial generated several hundred thousand free-text reports, we validated a large language model (LLM) to grade them.

## Methods

### Design and oversight

This was a stepped-wedge cluster randomized trial^10^ conducted in KPNC, an integrated health system serving more than 4 million members. Clusters were the 15 KPNC service areas. Two randomly selected service areas began the intervention in the first month, while the remainder crossed over to the intervention in a randomly assigned order until all were exposed (Figure 2). Fifteen service areas were randomized using random number generator software that determined the order of rollout. Exact rollout dates were determined by scheduled department meetings, which included a brief 20-minute education session about the CXR dashboard. Rollout dates were unevenly distributed but at a frequency of 1-2 service areas per month. A stepped-wedge design was chosen because the dashboard was to be deployed region-wide and a staged rollout was the only feasible way to train radiologists; the design also allows adjustment for secular trends. Radiologists could not be blinded. ED clinicians were not told when the radiologists in their service area had been trained, and outcome assessment was automated.

The KPNC Institutional Review Board approved the study (ID 2142507) with a waiver of informed consent. The trial is registered at ClinicalTrials.gov (NCT06785246). The protocol, outcomes, and analysis plan were specified in the funded proposal (2023) before enrollment.

### Participants

We included all STAT priority CXRs obtained during an ED or inpatient encounter at any KPNC facility and interpreted by a KPNC radiologist between June 19, 2024, and June 19, 2025. The unit of analysis was the CXR examination. A total of 521,641 STAT priority ED and inpatient radiograph reports and associated DICOM data were collected. We excluded same encounter follow-ups (114,103), non-CXR procedure codes (2,655), and exams missing patient location data (23), leaving 404,860 exams. There were no patient-level exclusions based on indication.

### Intervention

Grasshopper is a web application developed at KPNC that runs on the radiology workstation. When a radiologist opens an examination, the procedure code defines a reading context, and the dashboard queries the Epic-based EHR through web-service application programming interfaces. For CXRs, the module displays data organized around four categories of chest disease: infection or inflammation (e.g., white blood cell count, temperature), volume overload (e.g., BNP), neoplasia, and chronic lung disease, together with recent vital signs and excerpts of clinical notes (Figure 1). No radiologist action is required to retrieve the data. Before the trial, a beta version was used by about 5% of KPNC radiologists.

**Figure 1.**
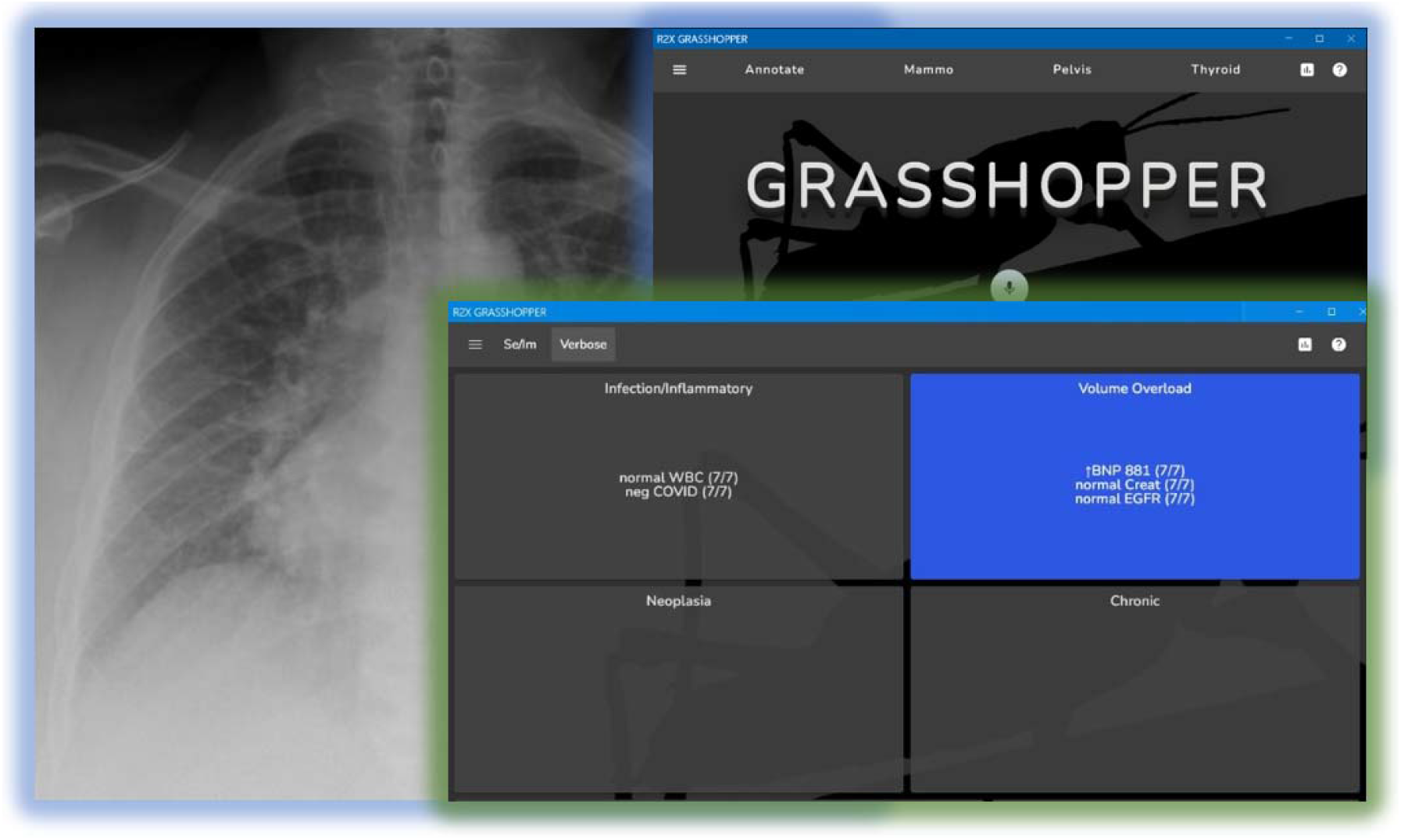
The Grasshopper chest radiograph module. De-identified screenshots of the dashboard (splash page, background; CXR module patient view, foreground) as displayed beside a CXR exam when a radiologist opens an ED chest radiograph, showing any pertinent available laboratory values, and vital signs. The dashboard does not cover diagnostic images in practice.

The intervention at each step consisted of activating the CXR module for radiologists in the service area and delivering education and promotion through department meetings and written materials. Department meetings included a 3-minute live introduction to the dashboard, a 7-minute educational video followed by a 5 to10-minute period for questions and discussion.

Training explained how specific markers can support or argue against a diagnosis (for example, favoring “pulmonary edema” over “opacities” when BNP is elevated). Use of the dashboard was encouraged but voluntary. Control periods reflected usual practice, in which radiologists had the order indication and could open the chart manually.

### Data sources

Report text, radiologist, reporting timestamps, and dashboard usage (whether it is open on the desktop) are data from a customized radiology database that captures metrics at report sign-off. ED discharge diagnoses, medication orders, and subsequent imaging came from the Epic Clarity database.

### Outcomes

The primary outcome was a specific report, defined as an impression that was either negative for acute disease (category 0) or abnormal with a single likely diagnosis for each finding (category 1). The comparator was a nonspecific abnormal report (category 2): one that used an unqualified nonspecific term such as “opacity” or “infiltrate,” offered more than one diagnosis for a finding, or used language of uncertainty such as “versus,” “not excluded,” or “correlate clinically” (Table 1). Impressions that made no statement about the lungs, mediastinum, or heart (category −1; for example, “as above” or a line-placement check) were excluded from report outcomes. We also examined the two components separately: negative versus any abnormal, and specific versus nonspecific among abnormal reports.

**Table 1.** Categories used by the large language model to grade chest radiograph report impressions.

| Category | Label | Criteria (abridged) | Example impressions |
| --- | --- | --- | --- |
| -1 | Non-categorical | No statement about the lungs, mediastinum, or cardiopulmonary findings; may refer only to the findings section. | "As above." "NG tube projects over the stomach." |
| 0 | Negative (specific) | Normal, or negative for acute or significant findings. | "No acute cardiopulmonary process." "Mild bibasilar atelectasis. No focal infiltrate." |
| 1 | Abnormal, specific | A single likely diagnosis for each finding. Likelihood terms (e.g., "consistent with," "suggestive of," "probable") are allowed. Uses terms that map to ICD-10 diagnoses (e.g., pneumonia, edema, effusion, pneumothorax, atelectasis, cardiomegaly). Every nonspecific term must be qualified by a specific diagnosis. | "Right upper lobe pneumonia. Small left pneumothorax." "Findings consistent with pulmonary edema." |
| 2 | Abnormal, nonspecific | More than one possible diagnosis for any finding; uncertainty terms (e.g., "versus," "or," "may," "not excluded," "correlate clinically"); or any nonspecific term (e.g., opacity, infiltrate, airspace disease, markings) not qualified by a diagnosis. Also any report not meeting another category. | "Right lung opacities." "Opacity suggestive of infection or malignancy." "Mild pulmonary edema or nonspecific infectious/inflammatory process." |

Secondary outcomes were (1) diagnostic concordance, defined among abnormal reports (category 1 or 2) as an impression that suggested the primary or any secondary ED discharge diagnosis; (2) a chest CT within 30 days after the CXR report was finalized; (3) an order for an antibiotic, a diuretic, either, or both within 7 days after the report was finalized; and (4) reporting duration in seconds, the time between the radiologist opening the study and signing off on the report.

### LLM classification of report impressions

Impressions were classified in a two-stage pipeline run in a secure institutional cloud environment (Databricks) through the health system’s LLM gateway, so that no report text left the organization. First, a regular-expression filter assigned category 0 to impressions matching common negative templates (e.g., “No acute cardiopulmonary process”). Second, each remaining impression was sent to the LLM (GPT-5.4, OpenAI^11^; reasoning effort set to “none”; maximum 4,000 output tokens) in a single prompt. The prompt instructed the model to act as an expert grader, supplied the category rules and 43 worked examples (Table 1), and required a JSON response containing the category and a brief explanation. Responses that could not be parsed were assigned category −1.

For diagnostic concordance, a second prompt gave the model the impression and the encounter’s discharge diagnoses, labeled primary or secondary. The model returned whether the impression suggested the primary diagnosis, a secondary diagnosis only, or neither. The prompt allowed reasonable clinical equivalence (for example, “airspace disease concerning for infection” matches pneumonia; “interstitial edema” matches congestive heart failure), directed that primary matches take precedence, and prohibited diagnoses not on the list. Reports in category 0 or −1 were not sent to the model for diagnosis concordance. Similar approaches have been used to structure free-text radiology reports.^12,13^ The initial analysis used GPT-4o; the full cohort was re-run with GPT-5.4 when it became available, and GPT-5.4 results are reported as the main analysis.

### LLM validation

We validated each task with a modified Delphi process^14^ (Figure 3). For specificity grading, 100 abnormal impressions were sampled at random. Two radiologists independently classified each as category 1 or 2, blinded to the model output and to each other. When the two disagreed, a third physician (emergency specialist), also blinded, broke the tie. A parallel process was applied to a second random sample of 100 abnormal reports for diagnosis matching. We set an a priori threshold of at least 85% agreement between the model and the expert consensus as the minimum acceptable performance.

### Radiologist survey

After the trial (April 29 to June 2, 2026), we sent an anonymous 6-item electronic survey to 375 current KPNC radiologists asking about years in practice, frequency of use of the CXR module in the past year, perceived value, and reasons for finding it helpful or unhelpful, with a free-text comment field.

### Statistical analysis

Analyses followed the intention-to-treat principle: each examination was assigned to control or intervention according to the scheduled rollout date of the service area in which it was read, regardless of whether the radiologist used the dashboard. Binary outcomes were analyzed with mixed-effects logistic regression and reporting duration with a linear mixed model.^15,16^ Models included a fixed effect for the intervention; fixed effects for time since study start, patient age, and radiograph view type; and random intercepts for radiologist, facility, and service area. We report adjusted odds ratios, model-standardized proportions for each condition, and adjusted absolute differences with 95% confidence intervals. Tests were two-sided with α=0.05. Secondary outcomes were not adjusted for multiplicity and should be interpreted as exploratory. Analyses used R, version 4.4.3.

## Results

### Cohort

The trial collected 521,641 radiographs interpreted by 387 radiologists across 15 service areas and 129 hospital locations. A total of 116,781 exams were excluded because of same encounter follow-ups (114,103), non-CXR procedure codes (2,655), and missing patient location data (23), leaving 404,860 ED and inpatient STAT priority CXRs: 256,303 (63%) in control periods and 148,557 (37%) in intervention periods (Figure 2). All 15 service areas crossed over as scheduled. After regular expression matching categorized 246,259 of 521,641 CXRs as negative (category 0), 275,382 reports were graded by the LLM (Table 2). After exclusions mentioned above, a total of 276,189 (69.5%) were negative, 50,058 (12.6%) were specific abnormal, and 70,868 (17.8%) were nonspecific abnormal. Thus, among the 120,926 abnormal reports, 59% were nonspecific.

**Figure 2.**
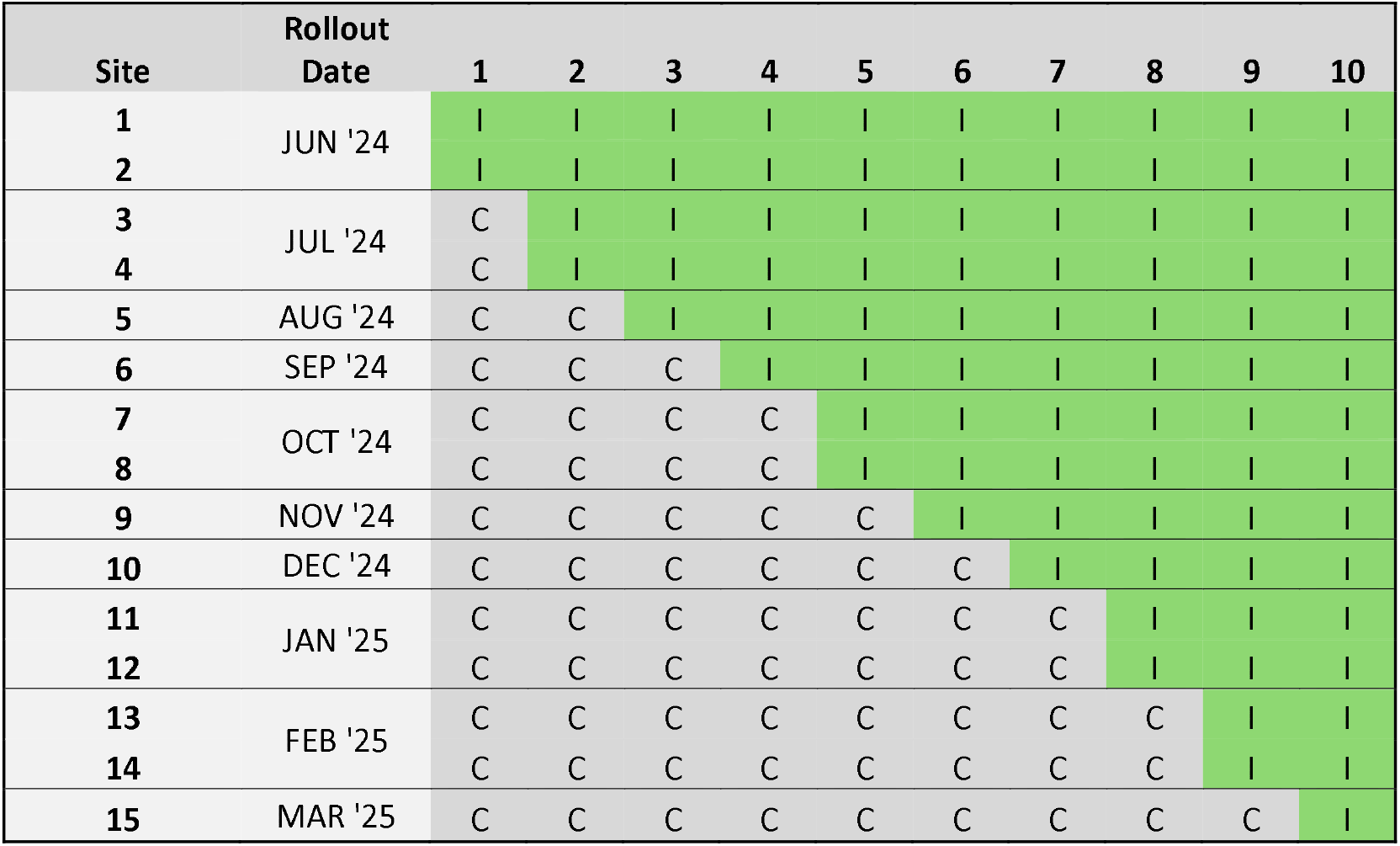
Stepped-wedge rollout. Rows are the 15 service areas in randomized order; columns are calendar months. Green cells labelled “I” indicate intervention periods, whereas grey cells labelled “C” represent control periods.

**Figure 3.**
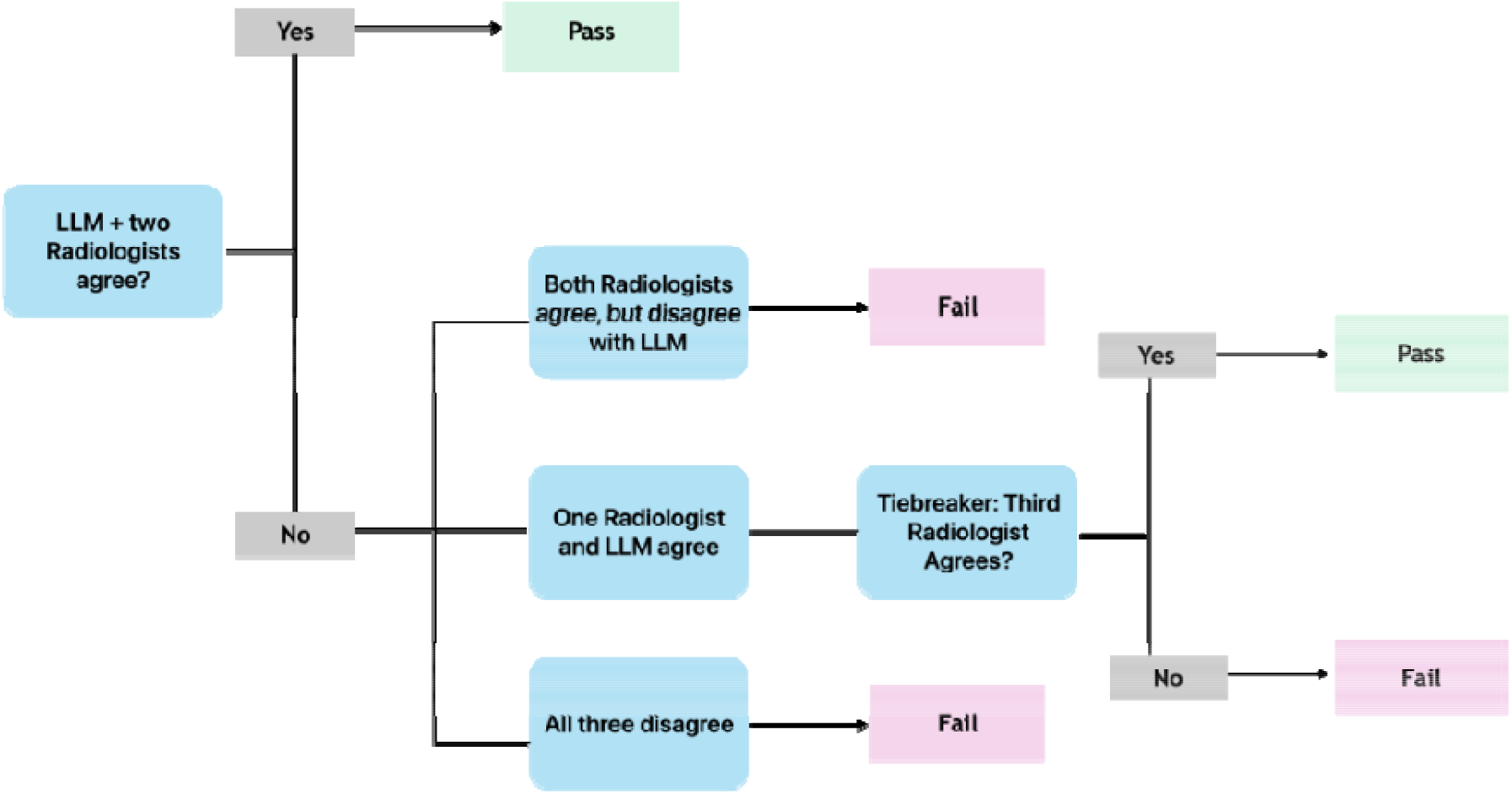
Delphi process for validating the large language model. Two blinded radiologists independently graded 100 randomly sampled abnormal impressions; a third blinded physician (emergency specialist) resolved disagreements; the consensus was compared with the model output against a prespecified 85% agreement threshold. The same process was applied to diagnosis matching.

**Table 2.** Examinations and report categories by study condition.

| Characteristic | Control periods | Intervention periods | Total |
| --- | --- | --- | --- |
| STAT priority chest radiographs, n (% of total) | 256,303 (63.3) | 148,557 (36.7) | 404,860 |
| Radiologists, n | 379 | 361 | 387 |
| Patient age, mean (SD), y | 57.6 (23.9) | 56.5 (24.4) | 57.2 (24.1) |
| Single-view, n (%) | 140,879 (55.0) | 76,809 (51.7) | 217,688 (53.8) |
| Two-view, n (%) | 114,247 (44.6) | 71,029 (47.8) | 185,276 (45.8) |
| Special view n (%) | 1,177 (0.5) | 719 (0.5) | 1,896 (0.5) |
| **Report category (GPT-5.4), n (% of all examinations)** |  |  |  |
| Non-categorical (−1), excluded from report outcomes | 4,865 (1.9) | 2,880 (1.9) | 7,745 (1.9) |
| Negative (0) | 175,272 (68.4) | 100,917 (67.9) | 276,189 (68.2) |
| Abnormal, specific (1) | 31,431 (12.3) | 18,627 (12.5) | 50,058 (12.4) |
| Abnormal, nonspecific (2) | 44,735 (17.5) | 26,133 (17.6) | 70,868 (17.5) |
| Abnormal reports (1 or 2) matched to a primary or secondary discharge diagnosis, n/N (%) | 30,453/76,166 (40.0) | 19,580/44,760 (43.7) | 50,033/120,926 (41.4) |

Report-category percentages use all examinations as the denominator. Radiologists are counted in each period in which they read at least one examination, so the period counts sum to more than the total. Among the 397,115 graded reports (categories 0–2), 69.5% were negative, 12.6% abnormal specific, and 17.8% abnormal nonspecific.

### LLM validation

With GPT-4o, both radiologists agreed with the model on specificity category in 83 of 100 impressions; the radiologists disagreed with each other in 10, and the third reader sided with the model in 6, giving 89% agreement with expert consensus. For diagnosis matching, initial agreement was 86%, and 6 of 8 tiebreaks favored the model, giving 92%. With GPT-5.4, agreement before adjudication was 84% for specificity (11 cases awaiting tiebreak at the time of this writing) and 89% for diagnosis matching (8 awaiting tiebreak); possible range for final agreement are 84-95% and 89-97%, respectively (Table 5).

### Primary outcome

The dashboard rollout did not change report specificity (Table 3). The adjusted proportion of specific reports was 82.2% (95% CI, 80.4% to 84.0%) in control periods and 82.1% (95% CI, 80.3% to 83.9%) in intervention periods, an adjusted difference of −0.10 percentage points (95% CI, −0.53 to 0.34; odds ratio [OR], 0.99; 95% CI, 0.96 to 1.03; P=.66). Neither component changed: the proportion of negative reports was 69.6% vs 69.5% (difference, −0.11 points; 95% CI, −0.63 to 0.41; P=.67), and among abnormal reports the proportion that were specific was 41.4% vs 41.3% (difference, −0.13 points; 95% CI, −1.14 to 0.87; P=.79).

**Table 3.**
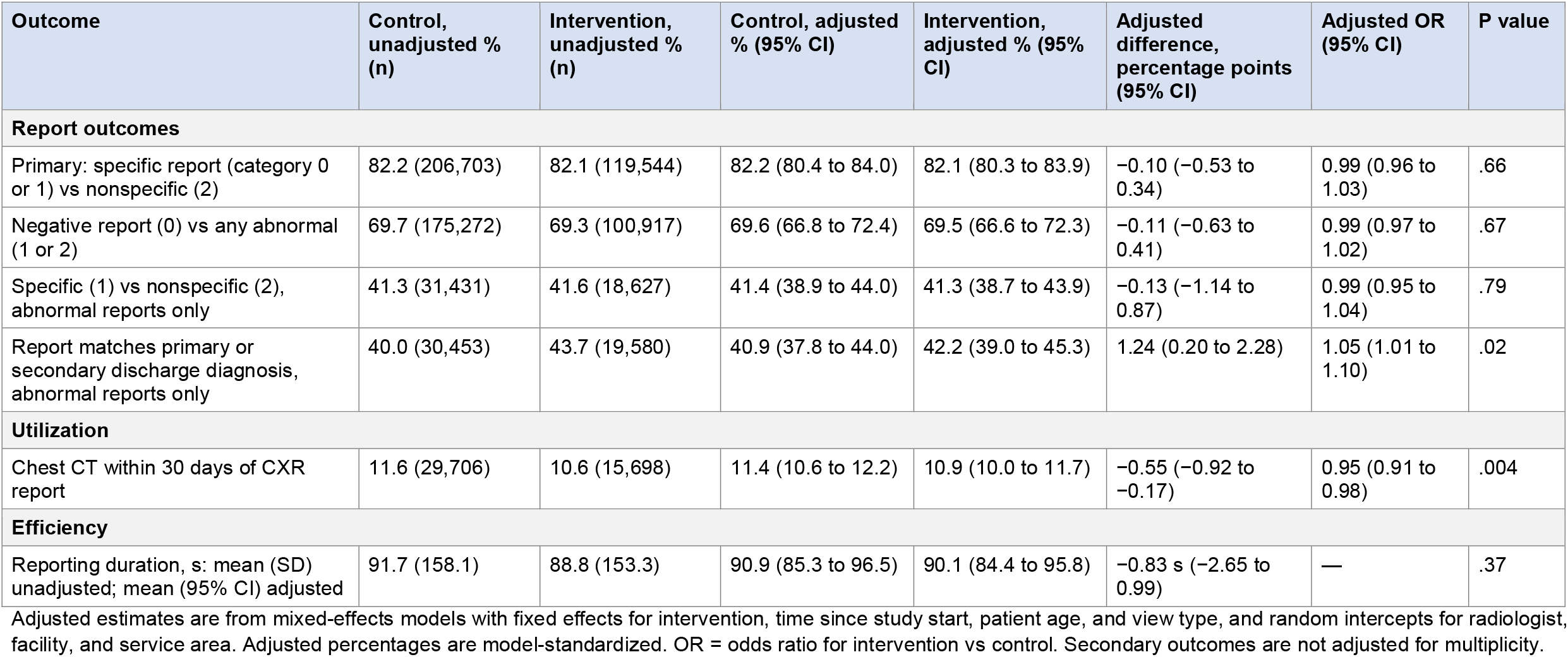
Primary and secondary outcomes (GPT-5.4 classification).

| Outcome | Control, unadjusted % (n) | Intervention, unadjusted % (n) | Control, adjusted % (95% CI) | Intervention, adjusted % (95% CI) | Adjusted difference, percentage points (95% CI) | Adjusted OR (95% CI) | P value |
| --- | --- | --- | --- | --- | --- | --- | --- |
| **Report outcomes** |  |  |  |  |  |  |  |
| Primary: specific report (category 0 or 1) vs nonspecific (2) | 82.2 (206,703) | 82.1 (119,544) | 82.2 (80.4 to 84.0) | 82.1 (80.3 to 83.9) | −0.10 (−0.53 to 0.34) | 0.99 (0.96 to 1.03) | .66 |
| Negative report (0) vs any abnormal (1 or 2) | 69.7 (175,272) | 69.3 (100,917) | 69.6 (66.8 to 72.4) | 69.5 (66.6 to 72.3) | −0.11 (−0.63 to 0.41) | 0.99 (0.97 to 1.02) | .67 |
| Specific (1) vs nonspecific (2), abnormal reports only | 41.3 (31,431) | 41.6 (18,627) | 41.4 (38.9 to 44.0) | 41.3 (38.7 to 43.9) | −0.13 (−1.14 to 0.87) | 0.99 (0.95 to 1.04) | .79 |
| Report matches primary or secondary discharge diagnosis, abnormal reports only | 40.0 (30,453) | 43.7 (19,580) | 40.9 (37.8 to 44.0) | 42.2 (39.0 to 45.3) | 1.24 (0.20 to 2.28) | 1.05 (1.01 to 1.10) | .02 |
| **Utilization** |  |  |  |  |  |  |  |
| Chest CT within 30 days of CXR report | 11.6 (29,706) | 10.6 (15,698) | 11.4 (10.6 to 12.2) | 10.9 (10.0 to 11.7) | −0.55 (−0.92 to −0.17) | 0.95 (0.91 to 0.98) | .004 |
| **Efficiency** |  |  |  |  |  |  |  |
| Reporting duration, s: mean (SD) unadjusted; mean (95% CI) adjusted | 91.7 (158.1) | 88.8 (153.3) | 90.9 (85.3 to 96.5) | 90.1 (84.4 to 95.8) | −0.83 s (−2.65 to 0.99) | — | .37 |

Adjusted estimates are from mixed-effects models with fixed effects for intervention, time since study start, patient age, and view type, and random intercepts for radiologist, facility, and service area. Adjusted percentages are model-standardized. OR = odds ratio for intervention vs control. Secondary outcomes are not adjusted for multiplicity.

### Secondary outcomes

Among abnormal reports, concordance between the impression and a primary or secondary discharge diagnosis was higher in intervention periods: 42.2% vs 40.9% (adjusted difference, 1.24 points; 95% CI, 0.20 to 2.28; OR, 1.05; 95% CI, 1.01 to 1.10; P=.02). The unadjusted proportions were 43.7% and 40.0%.

A chest CT was performed within 30 days after 10.9% of CXRs in intervention periods and 11.4% in control periods (adjusted difference, −0.55 points; 95% CI, −0.92 to −0.17; OR, 0.95; 95% CI, 0.91 to 0.98; P=.004). This corresponds to about 5.5 fewer chest CTs per 1,000 CXRs, or one fewer CT for every 180 CXRs read after rollout.

Prescribing did not change (Table 4). Within 7 days of the CXR report, antibiotics were ordered after 23.9% of CXRs in intervention periods vs 24.0% in control periods (adjusted difference, −0.09 points; 95% CI, −0.57 to 0.38; P=.70), and diuretics after 14.8% vs 14.7% (adjusted difference, 0.07 points; 95% CI, −0.33 to 0.46; P=.74). Orders for both an antibiotic and a diuretic were also unchanged (OR, 0.97; 95% CI, 0.93 to 1.02; P=.20). Results were similar at 24 and 48 hours (data not shown).

**Table 4.** Antibiotic and diuretic orders within 7 days after the chest radiograph report.

| Outcome | Control, unadjusted % (n) | Intervention, unadjusted % (n) | Control, adjusted % (95% CI) | Intervention, adjusted % (95% CI) | Adjusted difference, percentage points (95% CI) | Adjusted OR (95% CI) | P value |
| --- | --- | --- | --- | --- | --- | --- | --- |
| Antibiotic or diuretic | 30.2 (77,339) | 29.8 (44,211) | 30.0 (20.9 to 39.0) | 30.1 (21.1 to 39.2) | 0.19 (−0.30 to 0.69) | 1.01 (0.98 to 1.04) | .44 |
| Antibiotic and diuretic | 8.8 (22,460) | 8.6 (12,719) | 8.8 (5.1 to 12.4) | 8.6 (5.0 to 12.1) | −0.21 (−0.53 to 0.12) | 0.97 (0.93 to 1.02) | .20 |
| Antibiotic | 24.2 (61,920) | 23.6 (35,133) | 24.0 (15.9 to 32.1) | 23.9 (15.8 to 32.0) | −0.09 (−0.57 to 0.38) | 0.99 (0.96 to 1.02) | .70 |
| Diuretic | 14.8 (37,879) | 14.7 (21,797) | 14.7 (8.9 to 20.6) | 14.8 (8.9 to 20.6) | 0.07 (−0.33 to 0.46) | 1.01 (0.97 to 1.04) | .74 |

Adjusted estimates are from mixed-effects models specified as in Table 3.

**Table 5.** Agreement between the large language model and the blinded radiologist panel. The prespecified threshold for acceptable performance was ≥85%.

| Task (n=100 abnormal impressions each) | Model | Both radiologists agreed with model | Tiebreaks resolved in favor of model | Final agreement with expert consensus |
| --- | --- | --- | --- | --- |
| Specificity category (1 vs 2) | GPT-4o | 83 | 6 of 10 | 89% |
| Specificity category (1 vs 2) | GPT-5.4 | 84 | Tiebreak pending of 11 | Pending with range 84–95% |
| Match to discharge diagnosis | GPT-4o | 86 | 6 of 8 | 92% |
| Match to discharge diagnosis | GPT-5.4 | 89 | Tiebreak pending of 8 | Pending with range 89–97% |

Mean reporting duration was 90.1 seconds in intervention periods and 90.9 seconds in control periods (adjusted difference, −0.83 seconds; 95% CI, −2.65 to 0.99; P=.37).

### Radiologist survey

Twenty-eight radiologists responded (response rate: 28/375, 7.5%); 25 (89%) had more than 10 years in practice (Table 6). Reported use of the CXR module over the past year was split: 9 (32%) used it for more than half of CXRs, 5 (18%) used it sometimes, and 14 (50%) used it rarely or never. Sixteen (57%) rated the module very or slightly valuable, and 10 (36%) rated it not valuable. Among the 16 respondents who cited benefits, the most common were more specific reports (n=13), greater confidence (n=13), and greater efficiency (n=10). The most common reasons for not finding it helpful were “I do not think I need it” (n=9), insufficient screen space (n=4), and lack of time to try it (n=3). Free-text comments praised automatic retrieval of laboratory values (“knowing the BNP is elevated makes me more confident saying edema over atypical infection”) and cited intermittent loading failures, the module not opening automatically, and missing data when recent results were unavailable.

**Table 6.**
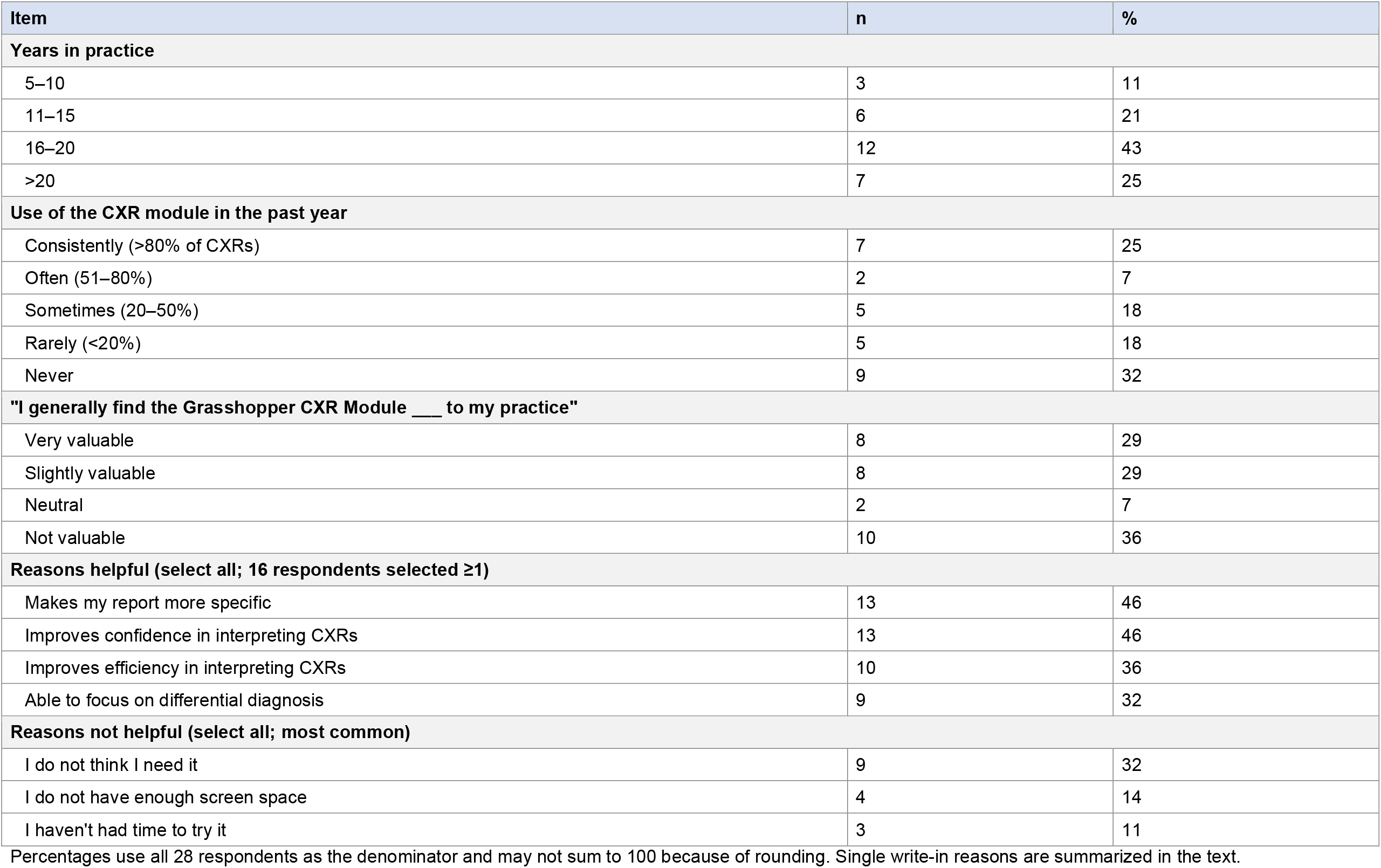
Radiologist survey responses (N=28).

## Discussion

In this stepped-wedge trial of 404,860 ED and inpatient STAT priority chest radiographs, rolling out an EHR-integrated dashboard with radiologist education did not change the specificity of report impressions. The confidence interval excludes any change larger than about half a percentage point, far smaller than the 1.2-point minimum difference the trial was designed to detect. Antibiotic and diuretic ordering was also unchanged. The rollout was associated with a 1.2-point increase in concordance between abnormal reports and the discharge diagnosis and a 0.55-point decrease in follow-up chest CT, with no increase in CXR reporting time.

### Why specificity did not change

Several explanations are plausible. First, the trial tested availability and promotion of the dashboard, not its use. Use was voluntary, and in our survey half of respondents reported using the module rarely or never, most often because they felt they did not need it. An intention-to-treat effect diluted by low uptake may be near zero even if the tool helps the radiologists who use it. Second, the data most likely to sharpen a diagnosis are often unavailable when an ED CXR is read: BNP and blood counts are frequently ordered at the same time as the radiograph and result afterward. Third, nonspecific language may reflect the inherent limitations of CXRs and radiologist hedging.^5,6^ The latter is influenced by radiologist training, medicolegal concern, and the real limits of the chest radiographs.^1^ A display of laboratory values may not shift those habits without structured reporting language or feedback. Finally, 70% of CXRs were negative and therefore already “specific,” leaving less room for improvement than the abnormal subset alone would suggest, although specificity did not change within the abnormal subset either.

### Secondary findings

The gain in diagnostic concordance without a change in specificity suggests that clinical context may influence which diagnosis a radiologist names more than whether the radiologist commits to one. Higher concordance could reflect better-informed interpretation, or simply alignment of the report with the clinician’s working diagnosis. The effect was small, the unadjusted difference (3.7 points) shrank by two thirds after adjustment for time and clustering.

The reduction in follow-up chest CT was statistically more robust and, given the volume of CXRs, could be meaningful at scale: about 5 to 6 fewer CTs per 1,000 CXRs. However, the mechanism is uncertain because the hypothesized mediator, report specificity, did not change. A report that better fits the clinical picture may leave less reason to order CT even when its wording is no more specific. Alternatively, the finding could reflect residual confounding by secular trends in CT use, to which stepped-wedge designs are vulnerable.^10,15^

Clinical information has been reported not to lengthen interpretation time^4^, and our data support that finding. Reporting time was unchanged, which addresses a common concern expressed by non-users in the survey.

### LLMs as a measurement instrument

Grading hundreds of thousands free-text reports by hand would not have been possible. A general purpose LLM, given written rules and examples but no fine-tuning, agreed with a blinded expert panel at a level we had set in advance as acceptable (GPT-4o); final validation of GPT-5.4 is pending adjudication of tiebreak cases. Agreement between the two radiologists themselves was imperfect (they disagreed on about 1 in 10 impressions), which places a ceiling on any grader. The Delphi framework offers a practical template for validating LLM-derived outcomes in pragmatic trials.

### Limitations

The trial took place in one integrated health system with a shared EHR and reporting platform, which may limit generalizability. Radiologists were not blinded, and although dashboard status was recorded at report sign-off, we did not analyze exam-level use and thus cannot estimate the effect of actually using the tool. The stepped-wedge design relies on correct modeling of time trends. Report specificity is a process measure; a specific report is not necessarily a correct one, and we did not assess diagnostic accuracy against a reference standard or patient outcomes such as return visits or missed diagnoses. The survey had few respondents and is subject to response bias.

### Implications

Making clinical data visible is not sufficient, on its own, to change how radiologists word CXR reports. Future work should test more active designs: surfacing data only when they discriminate between diagnoses, pairing the display with structured impression language, and giving radiologists feedback on their own specificity. Engagement should be logged for each examination so that the effect of use can be separated from the effect of availability. The measurement pipeline developed here can support those studies at low cost.

## Conclusions

Rollout of an EHR-integrated clinical dashboard for radiologists did not change the specificity of STAT priority chest radiograph reports or downstream antibiotic and diuretic prescribing. The dashboard rollout was associated with small improvements in report–diagnosis concordance and decreased follow-up chest CT use. The dashboard did not slow reporting workflow. Validated LLMs allowed report quality to be measured across an entire health system.

## Declarations

### Funding

This work was supported by The Permanente Medical Group (TPMG) Delivery Science and Applied Research program.

### Competing interests

Dr. Balkman developed the Grasshopper software as an employee of TPMG; the software is used internally at Kaiser Permanente and is not sold commercially. The other authors have no competing interests to declare.

### Ethics

The KPNC Institutional Review Board approved this study (ID 2142507) and waived informed consent. All relevant ethical guidelines were followed.

### Prior presentation

Portions of this work were accepted as an abstract to the Radiological Society of North America 2026 Annual Meeting (2026-SP-3491-RSNA).

### Use of AI tools

Large language models were used as a research instrument as described in the Methods.

### Data availability

Patient-level data contain protected health information and cannot be shared publicly; aggregate data are available from the corresponding author on reasonable request, subject to KPNC approval.

## Notes

### Competing Interest Statement

The authors have declared no competing interest.

### Clinical Trial

NCT06785246

### Author Declarations

The Institutional Review Board of Kaiser Permanente Northern California gave ethical approval for this work (IRB #2142507, approved March 13, 2024, expedited review, minimal risk). The Institutional Review Board of Kaiser Permanente Northern California, also serving as the Privacy Board, waived the requirements for informed consent (45 CFR 46.116) and HIPAA authorization (45 CFR 164.512(i)(1)(i)).

